# AI-based model for T1-weighted brain MRI diagnoses Amyotrophic Lateral Sclerosis

**DOI:** 10.1101/2024.04.26.24306438

**Authors:** Rosanna Turrisi, Federica Forzanini, Mario Stanziano, Anna Nigri, Davide Fedeli, Carrara Giovanna, Lequio Laura, Umberto Manera, Cristina Moglia, Maria Consuelo Valentini, Andrea Calvo, Adriano Chiò, Annalisa Barla

## Abstract

Amyotrophic Lateral Sclerosis (ALS) is an incurable deadly motor neuron disease that causes the gradual deterioration of nerve cells in the spinal cord and brain. It impacts voluntary limb control and can result in breathing impairment. ALS diagnosis is often challenging due to its symptoms overlapping with other medical conditions and many tests must be performed to rule out other conditions, as easily identifiable biomarkers are still lacking. In this study, we explore T1-weighted (T1w) brain Magnetic Resonance Imaging (MRI), a non-invasive neuroimaging approach which has shown to be a reliable biomarker in many medical fields. Nonetheless, current literature on ALS diagnosis fails to retrieve evidence on how to identify biomarkers from T1w MRI.

In this paper, we leverage Artificial Intelligence (AI) methods to unveil the unexplored potential of T1w brain MRI for distinguishing ALS patients from those who have similar symptoms but different diseases (*mimicking*). We consider a retrospective single-center dataset of brain T1-weighted MRIs collected from 2010 to 2018 recruited from the Piemonte and Valle d’Aosta ALS register (PARALS). The collection includes 548 patients with ALS and 106 with mimicking diseases. Our goal is to develop and validate a ML diagnostic model based exclusively on T1w MRI distinguishing the two classes. First, we extract a set of radiomic features and two sets of Deep Learning (DL)-based features from MRI scans. Then, using each representation, we train 8 binary classifiers. The best results were obtained by combining DL-based features with SVM classifier, reaching an F1-score of 0.91, and a Precision of 0.88, a Recall of 0.94, and an AUC of 0.7 considering the ALS group as the positive class in the testing set.

## 1. Introduction

Amyotrophic lateral sclerosis (ALS) is a fatal neurodegenerative condition characterized primarily by the progressive deterioration of upper motor neurons in the primary motor cortex and lower motor neurons in the brainstem and spinal cord. This degeneration leads to various manifestations in patients, such as muscle fiber atrophy, fasciculations, dysarthria, dysphagia, and a relentless progression towards paralysis, ultimately culminating in respiratory failure and death. The average life expectancy after diagnosis ranges from two to four years (Chiò et al., 2009).

The intricate nature and diverse manifestations of ALS contribute to the diagnostic challenges associated with the disease. Indeed, ALS can manifest in different phenotypes, encompassing not only motor symptoms but also non-motor symptoms like cognitive impairments and behavioral changes (Swinnen and Robberecht, 2014). This broad spectrum of presentations often results in clinical overlaps with other prevalent neurological disorders or conditions.

Compounding the diagnostic complexity is the absence of a specific biomarker that can reliably indicate the presence of ALS. Consequently, clinical diagnosis hinges on the exclusion of other disorders that may exhibit similar symptoms (Traynor et al., 2000; Yedavalli et al., 2018).

The challenge in diagnosing ALS often leads to a prolonged period of uncertainty, averaging about one year from the onset of symptoms to establish a definitive diagnosis (Falcão de Campos et al., 2022). Given the relatively short life expectancy of ALS patients, research into diagnostic biomarkers and accelerating the diagnostic process is crucial to promptly initiate interventions that can potentially improve patient outcomes and quality of life.

Among medical imaging tools for ALS diagnosis, Magnetic Resonance Imaging (MRI) stands out as a readily available and non-invasive approach, making it an ideal method for pinpointing radiological biomarkers (Plewes and Kucharczyk, 2012). Additionally, it holds immense promise as an imaging modality for developing automated diagnostic tools through Machine Learning (ML), which aims to create data-driven predictive models. In this context, T2-weighted MRI, Diffusion Tensor (DT) MRI and Fluid-Attenuated Inversion Recovery (FAIR) MRI are the preferred techniques employed by ALS specialists (Staff et al., 2015; Wang et al., 2011; Waragai, 1997). Although T1-weighted (T1w) MRI is widely used in the medical field for identifying subtle changes in tissue structure and abnormalities, only few studies have incorporated it in ALS research (Ignjatović et al., 2013; Ishaque et al., 2019; Schuster et al., 2016). This limited use of T1w MRI is attributed to the challenge that physicians face in extracting meaningful information from this examination, as well as the lack of large available datasets.

However, some studies suggest that T1w MRI may be a biomarker for ALS. Indeed, in (Ishaque et al., 2019), authors demonstrate that T1w MRI textures are associated with degenerative changes in the corticospinal tract in ALS patients. In (Nitert et al., 2022) the authors find that motor cortex neuroimaging parameters are more sensitive biomarkers. Still, to the best of our knowledge, current literature did not employ neither large datasets nor advanced statistical methods to fully explore the use of T1w MRI. Moving a step forward, this work investigates the potential of T1w images when analyzed by artificial intelligence methods and builds a classification task performing differential diagnosis, using the largest known corpus of T1w MRI of ALS patients.

Specifically, we firstly conduct a statistical study of the T1w MRI scans and the clinical variables, namely age and sex. Secondly, we extract relevant features from the images. This step is performed in two different ways. One approach relies on Radiomics, an emerging area of medical imaging providing a quantitative description of the image. The second approach uses a pre-trained Deep Learning (DL) model as feature extractor. Each MRI representation is finally used as input for a binary classifier. Eight different classifiers are trained and compared, using ML and DL approaches.

Results show that the best combination of MRI representation and ML model can reach an F1-score of 0.91, and a Precision of 0.88, a Recall of 0.94, and an AUC of 0.7, suggesting the T1w MRI as potential diagnostic biomarker for ALS.

## 2. Materials and Methods

In this section, we illustrate the characteristics and enrollment criteria of the ALS dataset and provide a detailed description of all steps of the analysis pipeline: feature extraction, classification algorithms, experimental design, and performance metrics.

### 2.1 Dataset

This retrospective single-center study is performed in accordance with the Declaration of Helsinki, after obtaining patients’ informed consent, and it has been conducted over a 8 year period, from 2010 to 2018 by the *Centro Regionale Esperto sulla SLA (CRESLA)* research center of the Department of Neurosciences of the University of Turin (Italy). The cohort consists of 654 subjects, of which 548 are affected by ALS and 106 present *mimicking* pathologies. The term *mimicking* refers to patients with an ALS-mimic disorder, not necessarily neurological, whose clinical presentation is very similar to that characterizing the onset of ALS. Specifically, mimicking patients were affected by cervical spondylotic myelopathy (n=62), myasthenia gravis (n=28), monomelic amyotrophy (Hirayama) (n=10), and peripheral motor neuropathy (n=6). All ALSmimic patients were initially recruited as suspected or possible ALS cases and were later diagnosed as mimicking.

As such, this binary classification problem is fairly unbalanced. To overcome this problem we synthetically obtain balance between classes by resorting to oversampling techniques that generate synthetic data in the minority class, i.e. mimicking cases.

For each patient, the dataset comprises two demographic features, which are the patient’s age at MRI scan and the biological sex of the patient, and a volumetric T1-weighted Magnetic Resonance Images (MRI) scan of the brain. Specifically, the MRI images were obtained by using a 1.5 Tesla scanner at an isotropic resolution of 1 mm, which means that the voxel size is 1 mm × 1mm × 1mm. The size of brain MRI images is 512×512×z voxels, thus z axial slices of dimension 512×512 were acquired for each patient. The number of brain axial slices is not the same for all patients, the mode being z = 120.

### 2.2 Feature extraction

Image classification relies on recognizing the underlying shapes and geometry within objects, encompassing tasks like image pre-processing through normalization, segmentation, feature extraction, and, ultimately, identification of the image content and semantic understanding. Here, we considered two distinct methods for feature extraction from T1w MRI scans. The first method capitalizes on Radiomics (Scapicchio et al., 2021), delving into the quantitative description of the image. The second approach utilizes a pre-trained Deep Learning (DL) (Goodfellow et al., 2016) model as a dedicated feature extractor.

#### 2.2.1 Radiomic features

Radiomics is an emerging and rapidly developing area of medical imaging. It provides a quantitative description of medical images, obtained with different non-invasive imaging modalities, through the high-throughput extraction and analysis of large amounts of reproducible features which represent textural information (Scapicchio et al., 2021).

For radiomic feature computation, we need to first segment the Volume of Interest (VOI), that is the brain region of interest, masking all non-brain tissues and the skull. This task poses significant challenges due to the close spatial proximity and similar intensities of non-brain tissues, especially in T1w images. To this aim, we employed a deep learning-based image segmentation approach by utilizing the open-source Python tool DeepBrain (Itzcovich, 2023). Specifically, DeepBrain relies on a pre-trained convolutional neural network (CNN) architecture based on the U-Net (Ronneberger et al., 2015) for efficient brain extraction from T1w MRI images.

Once the VOI was obtained, the radiomic features extraction was performed by means of PyRadiomics (van Griethuysen et al., 2017), an open source Python package developed for the computation of radiomic signatures of medical imaging data. The features extractor module of PyRadiomics library takes as input both the MRI image and segmented VOI mask. This extractor was customized to normalize and resample the image and the mask if necessary and to check the validity of VOI defined in the mask. Finally, we extracted 107 radiomic features, including 18 features of First Order Statistics, 14 shape-based features, 24 Grey Level Co-occurrence Matrix features, 14 Gray Level Dependence Matrix features, 16 Grey Level Run Length Matrix features, 16 Gray Level Size Zone Matrix features, and 5 Neighboring Gray Tone Difference Matrix features.

#### 2.2.2 Deep Learning-based features

Convolutional neural networks (CNNs) (Lecun and Bengio, 1995) have been revolutionary in extracting meaningful features from images, as they can automatically find spatial-related features at different levels of image representation. Indeed, the first convolutional layers learn basic features, such as object shapes, while deeper layers grasp abstract and high-level features. The feedforward layer, performing the classification, is instead tailored to the specific task on which the model is trained. With the advent of available models pre-trained on large datasets, several computer vision applications rely on using the convolutional layers as feature extractor (Dara and Tumma, 2018; Jogin et al., 2018; Yildirim et al., 2019; Zhao and Du, 2016). Although there is a vast choice of pre-trained models on 2D images, only few models pre-trained on 3D medical images are publicly available.

In this work, we adopted a pre-trained 3D-CNN originally proposed in (Turrisi et al., 2023). The architecture consists of 8 convolutional layers followed by a feedforward layer and it has been trained on T1w volumetric brain MRI scans to distinguish between healthy subjects and patients with Alzheimer’s Disease. Although the model has been trained for a distinct clinical purpose, the model’s training data align closely with the dataset utilized in this manuscript, making this pre-trained model remarkably suitable for feature extraction in this context.

Hence, for each MRI, we extracted two deep layers from the 3D-CNN: the last convolutional layer (DL8) and the penultimate one (DL7), see Figure 1. Both DL representations consist of 256 features.

**Figure 1:**
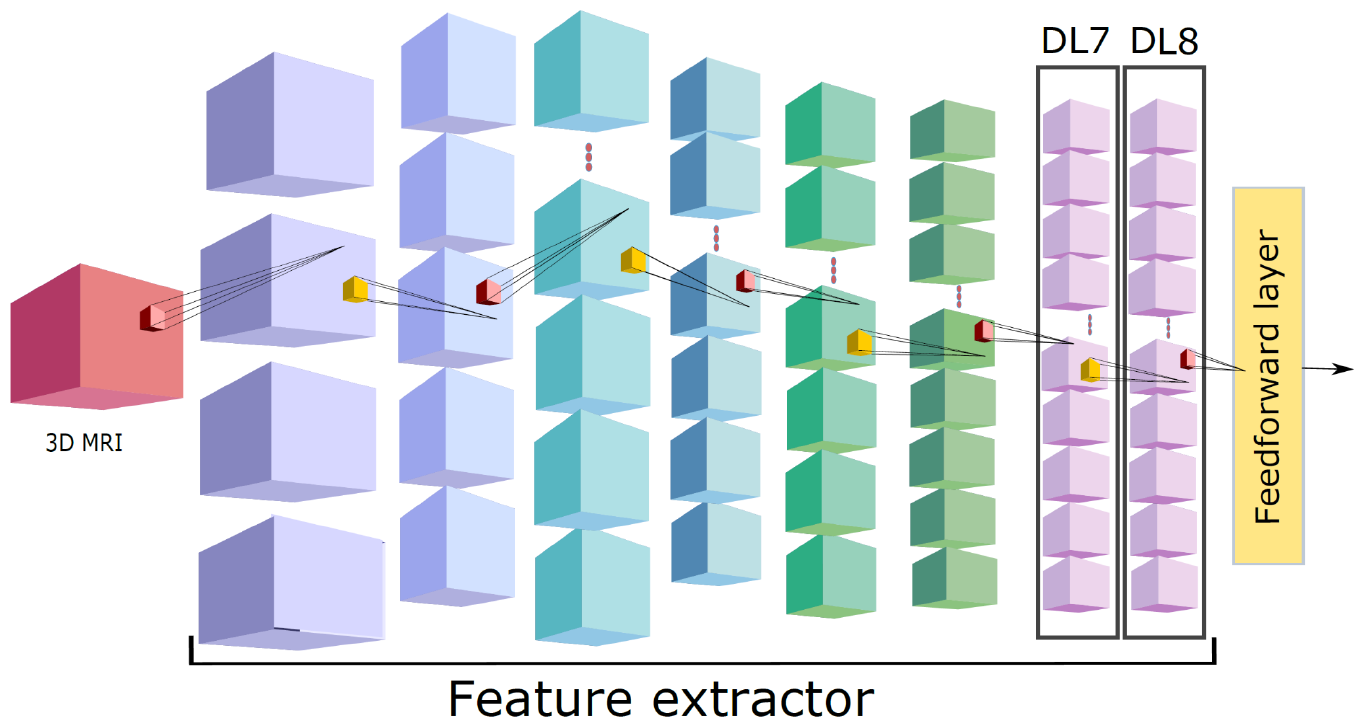
The pre-trained model is used as a feature extractor. ALS and *mimicking* brain images are provided as input to extract the last (DL8) and penultimate (DL7) convolutional layer.

### 2.3 Classification algorithms

Once extracting the MRI representations, we used each feature vector as input for a classifier to discriminate between ALS and mimicking patients. In order to solve the binary classification task, we employed K-nearest neighbors (KNN), Random Forest (RF), and Support Vector Machine (SVM) classifiers (Hastie et al., 2009) with four different kernels: linear (L), Gaussian (RBF), polynomial (P) and sigmoid (S). We refer to these classifiers as ML classifiers. Also, we resort to DL training a Deep Neural Network (DNN) with two early stopping criteria: one on training (DNNt) and one on validation (DNNv), respectively (Goodfellow et al., 2016).

### 2.4 Experimental design and resampling strategies

For the ML classifiers, grid search has been performed to find the best hyper-parameters. In particular, we employed stratified-3-fold cross-validation in which the dataset has been shuffled with random state 42 and test size set to 20%.

For KNN we explored the number of nearest neighbors *k* varying among 3, 5, and 7.

In the Random Forest classifier, we searched for the best number of trees *N* among 5, 100, 200, 500.

Finally, for SVM classifiers, we searched for the optimal regularization parameter *C* among 0.0001, 0.001, 0.01, 0.1, 1, 10. Further for SVM with the Gaussian kernel we looked for the best kernel coefficient *γ* among 0.001, 0.01, 0.1, 1, 10 and for the polynomial kernel we investigated the polynomial degree *d* equal to 2 and 3.

Table 1 reports the found best hyper-parameters for each ML method and MRI representation.

**Table 1:** Optimal hyper-parameters found via grid search, for each method and image representation.

|  | Knn | RF | SVM (L) | SVM (P) |  | SVM (RBF) |  | SVM (S) |
| --- | --- | --- | --- | --- | --- | --- | --- | --- |
| | $k$ | $N$ | $C$ | $C$ | $d$ | $C$ | $\gamma$ | $C$ |
| Radiomics | 7 | 500 | 0.001 | 1.0 | 3 | 1.00 | 0.01 | 0.001 |
| DL7 | 7 | 200 | 0.001 | 0.1 | 3 | 1.0 | 0.1 | 0.0001 |
| DL8 | 7 | 200 | 0.01 | 1.0 | 2 | 10 | 0.001 | 0.0001 |

**Table 1:** Patient Characteristics. Data are numbers of patients with percentages in parentheses.

| Variable | ALS | Mimicking |
| --- | --- | --- |
| **Sex** |  |  |
| F | 230 (42.0) | 34 (32.1) |
| M | 318 (58.0) | 72 (67.9) |
| **Age** |  |  |
| <60 | 171 (31.1) | 42 (39.6) |
| 60-75 | 293 (53.6) | 50 (47.2) |
| >75 | 84 (15.3) | 14 (13.2) |

With DL, we considered a network of 3 layers with 50, 3,2 neurons, respectively. Dropout with a probability of 0.2 was applied after each layer. The network underwent training with the objective of minimizing the cross-entropy loss function, employing the Adam optimizer with a learning rate set to 0.001. Additionally, an l2-penalty equal to 0.001 was incorporated. We set a maximum of 200 epochs with a patience of 50 for early stopping. Early stopping was implemented in two ways. In the first scenario, acknowledging the limited sample size, we aimed to preserve the training set size. Consequently, we did not employ an additional validation set and executed early stopping based on the F1-score of the training set. We refer to this method as DNNt. In the second approach, we utilized 20% of the training set as a validation set to enhance model generalization. This method is referred to as DNNv. To assess the stability with respect to weights initialization, we ran each DNN model on 5 trials.

Once fixed the hyper-parameters, we compared the performance of the adopted models for each MRI representation. In order to guarantee a correct assessment of model performance and stability, we set up a stratified-nested K-fold cross-validation loop with K=3. Hence, the dataset has been split 3 times into training (n=436) and testing (n=218) set by keeping the same subject group proportions. Further, to overcome the unbalanced classes problem, we augmented the training set by using Synthetic Minority Oversampling Technique (SMOTE) technique (Chawla et al., 2002). SMOTE is a sophisticated oversampling method, whose main idea is to generate synthetic examples of the minority class within the convex hull of the all minority class instances in the training set. More specifically, a random sample (S1) from the minority class is first chosen, then *k=5* samples of the nearest neighbors are identified. After randomly selecting one of them (S2), a line between S1 and S2 is drawn and a synthetic example is created at a randomly selected point within the line. This approach can significantly improve learning procedure, allowing the classifier to generalize better since it can learn larger and less specific decision regions for the minority class examples. The final size of the augmented training set was n=730.

### 2.5 Performance metrics

To evaluate the quality of learning algorithms, the predictions of the learned classifiers were compared to the true classes of test data by computing performance measures and the confusion matrix. In order to provide comprehensive assessments of the learning problems we considered Precision, Recall, F1-score, AUC and AUPRC (Hastie et al., 2009). All metrics are available in the Scikit-learn library of Python (Fabian et al., 2011).

## 3. Results

### 3.1 Data exploration

#### 3.1.1 Patient Demographic Characteristics

As reported in Table 1, the dataset consists of 548 subjects (M=318, F=230; mean age=64±12 years) affected by ALS and 106 ALS-mimic subjects (M=72, F=34; mean age=60±15 years). The median age of ALS and mimicking patients are 65 and 62, respectively. The youngest and oldest ALS patients are 20 and 95 years old, while the age of mimicking patients ranges from 17 to 85.

Figure 2, which compares the quantiles of the age distribution of patients and a set of theoretical quantiles from the Normal probability distribution, suggests that the age distributions of both ALS and ALS-mimic patients are tailed. Specifically, they are mainly skewed left with respect to the Normal distribution which means that there is more data to the left of the theoretical distribution, since the quantiles are at much lower values than they would be in a Normal distribution.

**Figure 2:**
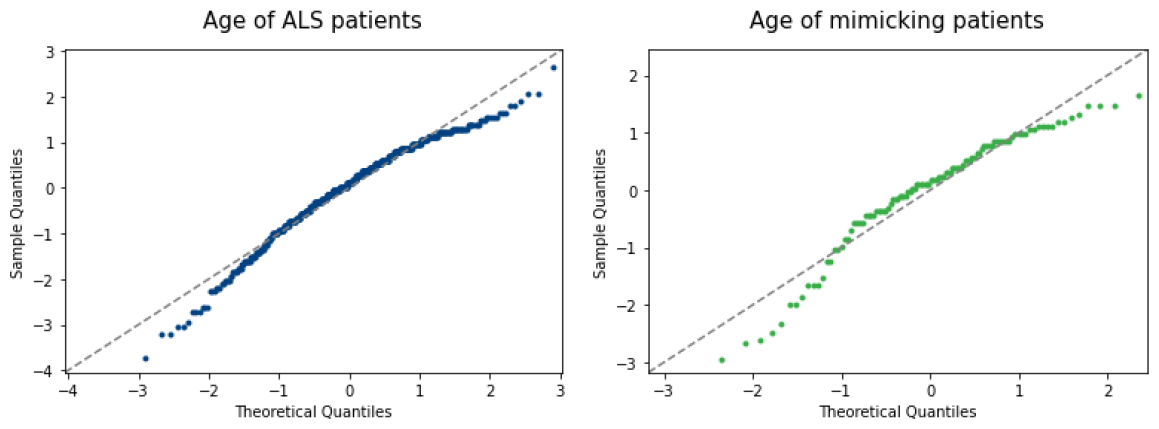
Comparison between quantile-quantile plot related to age distribution of ALS (blue dots, left panel) and ALS-mimics (green dots, right panel) patients. The dashed gray line is the reference line that shows where the points would fall if the data were normally distributed.

Statistical tests were performed in order to assess the inter-group differences between ALS and mimicking patients. In particular the two-sample Kolmogorov-Smirnov test was used to compare the underlying continuous distributions of the patients’ age in the two groups. Similarly, Pearson’s χ2 test was applied to the biological sex feature. Table 2 shows the values of the statistic and the p-value obtained from the two statistical tests. They indicate that there are no statistically significant inter-group differences between ALS and ALS-mimic patients in terms of age distribution and sex frequency, since both the p-values are greater than the fixed threshold of 0.05. The results obtained from the analysis of the distributions of the demographic features suggest that patients with mimicking disorders match for age and sex those with ALS.

**Table 2:** Statistical test verifying the inter-group difference for ALS and mimicking patients with respect to biological sex and age.

| Variable | Statistical Test | Results | p-value |
| --- | --- | --- | --- |
| **Sex** | Pearson's $\chi^2$ | 3.34 | $0.07 > 0.05$ |
| **Age** | Kolmogorov-Smirnov | 0.11 | $0.29 > 0.05$ |

#### 3.1.2 MRI Characterization

We conducted an analysis of the signal intensity of the MRI scans in terms of gray level of voxels by studying the frequency distribution of the voxel values. For each MRI image, we computed an histogram by considering a fixed discretization (bin width) of the voxel value range. Voxels with null value were excluded since they correspond to the background. Then, the mean frequency distributions of gray level of voxels per group were computed by averaging the histograms obtained from the MRI images of ALS patients and those of mimicking patients, respectively. The resulting two histograms are shown in Figure 3. These plots illustrate how the MRI images of the two groups have almost the same mean frequency distribution of voxel values. In both cases, the histogram extends to large voxel values with small voxel counts. This implies that only a few voxels are very bright. On the other hand, there is a huge amount of voxels with a value of less than 60, which means that most voxels are very dark.

**Figure 3:**
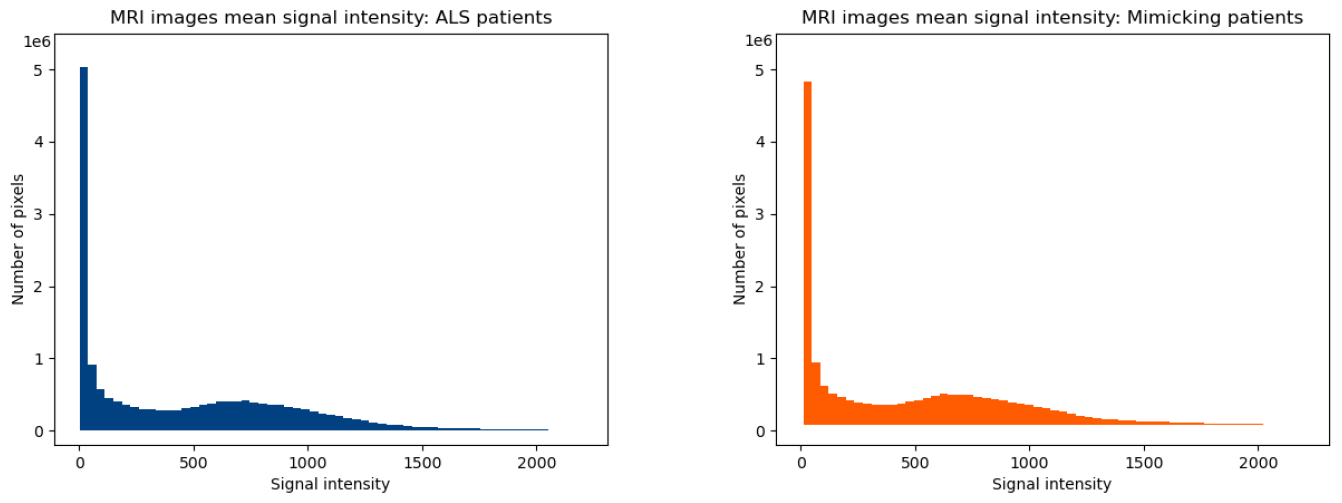
Results of MRI signal analysis. Comparison between the mean frequency distribution of voxel values in MRI images of ALS patients (blue histogram on the left) and mimicking patients (orange histogram on the right).

### 3.2 ALS differential diagnosis

In Table 3 we report the average and standard deviation over the 3 splits for the F1-score computed on the testing sets for the previously described classifiers, given one of the three feature representations (Radiomics, DL7, DL8) as input. Note that results of the DNNs are also averaged also on the 5 trials. The best performing model is the one combining the DL7 representation and SVM (P), with an average F1-score of ∼92%. In the following, we dig into the best classification results for each representation. In particular, we report Precision, Recall, F1-score, area under the receiver operating characteristic curve (AUC) and the area under the precision-recall curve (AUPRC) for ALS and mimicking classes (Table 4).

**Table 3:** Mean values and standard deviations for the F1-score (%) index evaluated for different combinations of feature type and Machine Learning method.

| Methods | Features |  |  |
| --- | --- | --- | --- |
|  | Radiomics | DL7 | DL8 |
| **SVM (L)** | 85.10 ± 0.73 | 90.35 ± 1.17 | 88.04 ± 1.40 |
| **SVM (P)** | 85.82 ± 2.29 | **91.76 ± 0.31** | **89.77 ± 2.16** |
| **SVM (RBF)** | 83.84 ± 2.90 | 87.71 ± 0.82 | 85.92 ± 1.12 |
| **SVM (S)** | 78.55 ± 8.36 | 91.11 ± 0.62 | 80.65 ± 4.26 |
| **Knn** | 68.99 ± 5.13 | 70.15 ± 3.11 | 77.65 ± 1.23 |
| **RF** | 85.92 ± 3.45 | 90.58 ± 0.46 | 86.97 ± 0.68 |
| **DNNt** | 85.85 ± 1.23 | 90.11 ± 1.07 | 85.85 ± 2.07 |
| **DNNv** | **86.83 ± 2.72** | 88.45 ± 1.91 | 86.53 ± 2.49 |

**Table 4:** Evaluation of the best model for each MRI representation, in terms of F1-score, precision, recall, AUC and AUPRC.

|  | Features and Best Classifier |  |  |
| --- | --- | --- | --- |
| Metrics | Radiomics + DNNv | DL7 + SVM (P) | DL8 + SVM(P) |
| F1-score | 0.87 | 0.92 | 0.90 |
| Precision | 0.89 | 0.88 | 0.87 |
| Recall | 0.85 | 0.95 | 0.92 |
| AUC | 0.31 | 0.69 | 0.64 |
| AUPRC | 0.36 | 0.91 | 0.87 |

The best performance, obtained by the combination of the DL7 representation and an SVM classifier with a polynomial kernel, yields high scores for all metrics. Specifically, it achieves a precision of 0.88, recall of 0.96, AUC of 0.69, and AUPRC of 0.91.

Similarly, in Figure 4, we display the confusion matrices for the best classifiers for each representation.

**Figure 4.**
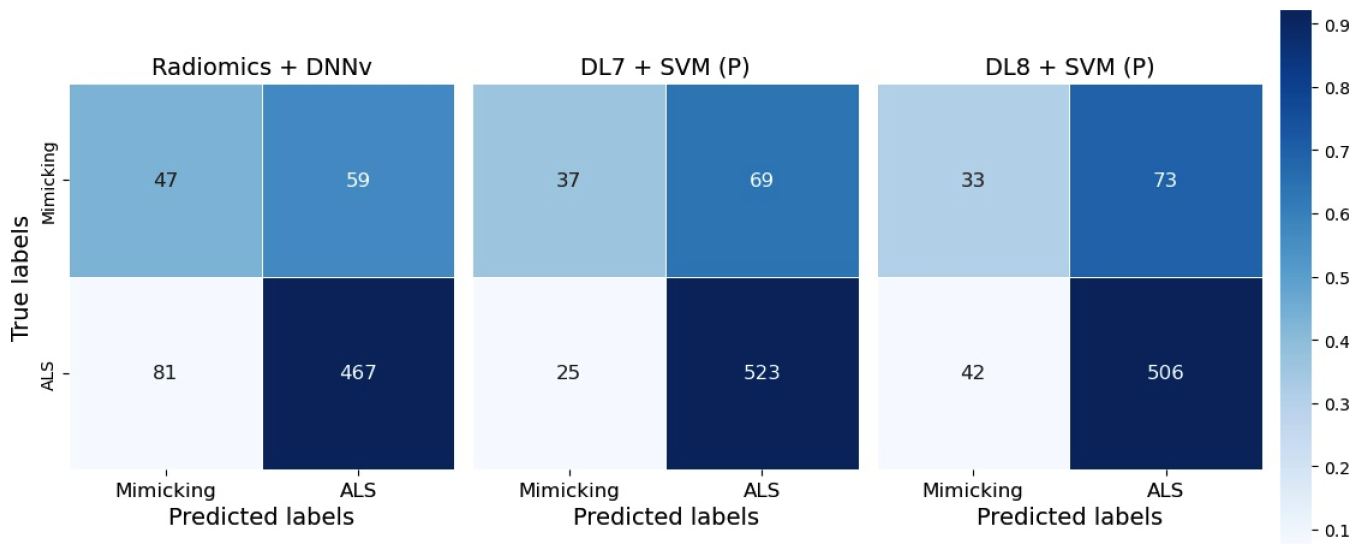
Confusion matrix of the best model for Radiomics+DNNv, DL7+SVM (P) and DL8+SVM (P) features.

## 4. Discussion and Conclusion

This study explores the potential of utilizing ML algorithms to analyze T1w MRI, recognizing that this imaging procedure is not presently incorporated into standard clinical protocols, as physicians currently cannot derive diagnostic information from this MRI modality. Indeed, Figure 3 reports a preliminary analysis on the overall T1w MRI intensity in the two classes underlining that ALS and mimicking distributions are indeed very similar. It is therefore unlikely that statistical methods based on single-features will be sufficient to reach acceptable classification performance.

Therefore, we argue that this MRI modality calls for more advanced ML and DL methods of feature representation and learning. Following this avenue, we here show that very high performance metrics can be reached in various configurations.

We considered eight different binary classifiers and three data representation approaches, and compared the classification performance based on several metrics.

The best result is achieved by using the penultimate convolutional layer (DL7) of the deep neural network, discussed in Section 2.2.2, as input of a Support Vector Machine endowed with a polynomial kernel of degree 3. This configuration reaches a F1-score of 91.76 ± 0.31, averaged over the k=3 fold of a cross-validation loop, see Table 3. High performance is further confirmed by additional metrics (Table 4). Indeed, this pipeline achieves a precision of 0.88, recall of 0.96, AUC of 0.69, and AUPRC of 0.91. If we consider the alternative representations, i.e. the last convolutional layer in the same deep neural network (DL8) or the engineered radiomic representation, we still manage to reach very good prediction F1-score: 89.77 ± 2.16 and 86.83 ± 2.72, respectively. The lower value associated with the radiomics features is in line with current literature (Avanzo et al., 2020; Parekh and Jacobs, 2019), where experts agree on the advantage of adopting DL-based approaches when the main goal is to maximize the prediction performance.

When using DL to represent data, we observe that classifiers using DL7 as a representational layer always outperform their counterparts using DL8, with the exception of Knn. This result is in line with (Sun et al., 2022), where the authors assert that using the penultimate layer’s feature is better than using the projection head, when dealing with out-of-distribution data. Here, we are exploiting the deep neural network described in Section 2.2.2 and trained on Alzheimer’s T1w MRI scans as a tool for feature extraction on the ALS T1w MRI exams, indeed considering an out-of-distribution dataset.

On average, all configurations lead to a median F1-score of 86. This corroborates that T1w is indeed informative to address the ALS vs mimicking problem, and that the key tool to achieve such results is the adoption of advanced ML and DL techniques which may capture more complex patterns that link the input variables to the clinical outcome. Limitations of this work are related to three main factors: unbalancedness of the two classes, partial exploration of DL configurations, and interpretability of results from DL approaches. To balance the mimicking class, we are actively working to expand and diversify the dataset to enhance the model’s robustness and generalization capabilities.

We did not have the option of choosing many alternatives for the deep neural network used for representation, as, to the best of our knowledge, there are no models trained on ALS T1w MRI scans. To improve this part of the pipeline, we plan to fine-tune the 3D CNN of Section 2.2.2 on the ALS differential diagnosis task. To address the issue of interpretability, our future work will focus on implementing attention models to automatically detect and highlight important regions within the images. This approach aims to enhance the transparency of our model’s decision-making process, providing clinicians with more confidence in its outputs. Additionally, we recognize the potential benefits of segmenting the brain and applying radiomics and deep learning features to assess performance in specific brain areas. This targeted analysis could offer valuable insights into the model’s strengths and weaknesses, paving the way for personalized diagnostics and treatment planning.

## Data Availability

All codes produced in the present study are available at https://github.com/rturrisige/T1wMRI4ALS_diagnosis

## Acknowledgments

The research project was mainly funded by the DECIPHER-ASL – Bando PRIN 2017 grant (2017SNW5MB - Ministry of University and Research, Italy). This work is partially funded by the European Union - NextGenerationEU and by the Ministry of University and Research (MUR), National Recovery and Resilience Plan (NRRP), Mission 4, Component 2, Investment 1.5, project “RAISE - Robotics and AI for Socio-economic Empowerment” (ECS00000035) as A. Barla is part of the RAISE Innovation Ecosystem.

